# Service Provider Competence, Motivation, and Resource Availability as Determinants of Quality Healthcare Delivery: A Cross-Sectional Study at Makeni Regional Hospital, Sierra Leone

**DOI:** 10.64898/2026.09.10.26362765

**Authors:** Aiah Lebbie, Abraham I. Jimmy, Sallu N. Kamara, Lee P. Gary

## Abstract

Quality healthcare delivery in low- and middle-income countries remains constrained by workforce and resource limitations. This study assessed service provider competence, service provider motivation, and resource availability as determinants of quality healthcare delivery at Makeni Regional Hospital in Northern Sierra Leone, from the service providers’ own perspective.

A cross-sectional study, descriptive in type, was conducted among 155 service providers, with sample size determined using the Yamane formula. Structured questionnaires were administered and analyzed using STATA to assess the relationship between competence, motivation, resource availability, and quality healthcare delivery. Most service providers (74.8%) reported possessing the required professional skills, and 91.6% perceived that competent staff resulted in improved delivery of quality healthcare; 94.8% associated competent personnel with improved patient outcomes. Motivation levels were markedly lower: only 7.1% of respondents were highly motivated, 16.1% were motivated, and 40.65% were somewhat motivated, while all respondents (100%) agreed that motivation influences quality of care delivery.

Resource availability showed the strongest association with quality outcomes, with 99.4% of respondents affirming that adequate resources affect quality healthcare delivery and 95.5% affirming that adequate resources result in improved delivery of quality healthcare; gaps were noted in bed capacity (45.8% reporting inadequacy) and equipment. Service provider competence, motivation, and resource availability were all found to significantly determine the delivery of quality healthcare at Makeni Regional Hospital, with motivation representing the weakest of the three despite high levels of provider competence.

Continuous professional development, formal motivation policies, and improved resource allocation are recommended to strengthen quality healthcare delivery.

## INTRODUCTION

Quality healthcare service encompasses the scope of medical services provided, staff clinical competence, hospital amenities, physician expertise, hospital ambiance, staff behavior, in-patient experience, and, most importantly, patient satisfaction (Kavya, 2019). The World Health Organization defines quality of care as the degree to which health services for individuals and populations promote desired health outcomes and are equitable, timely, effective, safe, and people-centered (UNFPA Sierra Leone, 2018). The Institute of Medicine similarly frames quality as the extent to which health services increase the likelihood of desired health outcomes and remain consistent with current professional knowledge, identifying safety, patient experience, effectiveness, efficiency, equity, and timeliness as its core dimensions (Atkinson et al., 2010).

Despite decades of accumulated global expertise on improving healthcare standards, policymakers in both high- and low-income countries continue to struggle to identify which quality-focused policies most improve health system outcomes, and significant variation in care standards persists even where systems are well resourced (Obina, 2019). This gap is most pronounced in Africa, which carries roughly 25% of the global disease burden yet holds only about 3% of the world’s health workforce; the continent’s health-worker density is estimated at 2.3 per 1,000 population, compared with 24.8 per 1,000 in the Americas (Alhassan et al., 2013). Human resources for health are widely regarded as the most valuable, and most fragile, input into any health system (Munanye, 2014).

Sierra Leone illustrates these constraints acutely. The country ranked fourth from last among 49 prioritized low- and middle-income countries in skilled health worker density as of 2010, and the 2014-2015 Ebola Virus Disease outbreak claimed the lives of 221 healthcare workers, further weakening an already fragile workforce (Mózo, 2018). Beyond staffing numbers, low provider motivation has been linked to patient irritability, absenteeism, long waiting times, informal fee charging, and labour unrest (Alhassan et al., 2013), while providers working in rural Sierra Leone report unfavorable working conditions, limited access to training, long working hours due to staff shortages, and restricted income-generating opportunities as key demotivating factors (Wurie et al., 2016).

The Government of Sierra Leone introduced the Free Health Care Initiative (FHCI) in 2010 to remove user fees for pregnant women, nursing mothers, and children under five, in response to persistently high maternal and child mortality (Witter et al., 2016). Even after the FHCI, Sierra Leone recorded the world’s highest maternal mortality ratio, at 1,360 deaths per 100,000 live births in 2015 (WHO, 2021), and fell short of the MDG4 and MDG5 targets of 450 maternal deaths per 100,000 births and 95 infant deaths per 1,000 live births (Jalloh et al., 2019). National survey data show a 2018 public perception of long wait times and widespread informal payment for care, disproportionately affecting poorer and less-educated citizens (Sanny, 2020), while Demographic and Health Survey trends show under-5, infant, and neonatal mortality falling between 2013 and 2019 but remaining well above global benchmarks (DHS, 2019).

## OUR MOTIVATION

Against this backdrop, this study investigated the factors affecting the delivery of quality healthcare at Makeni Regional Hospital, a referral hospital serving approximately 1.7 million people in Northern Sierra Leone, from the perspective of its own service providers.

Our specific objectives were to assess whether service provider competence, service provider motivation, and the availability of resources affect the delivery of quality healthcare at the hospital.

## METHODOLOGY

### Study Design and Setting

This was a cross-sectional, descriptive study conducted at Makeni Regional Hospital, a referral hospital in Makeni, the largest city in Sierra Leone’s Northern Province. The hospital serves residents of Bombali District, including Makeni itself, as well as patients referred from district hospitals across the Northern and North-West Provinces.

### Study Population and Sample

The study population comprised 210 clinical and administrative staff providing services at the hospital. The sample size for the quantitative component was determined using the Yamane formula at a 95% confidence level and 0.05 margin of error, yielding 138 respondents, which was increased to 155 to account for a projected 12% non-response rate. Proportional sampling, a form of stratified sampling, was used to draw respondents across the different cadres of healthcare providers in proportion to their representation in the study population. Providers who presented for duty during the data collection period and consented to participate were recruited.

### Data Collection

Primary quantitative data were collected using a structured questionnaire administered by trained data collectors over a three-week period, from 5 to 23 December 2022, across morning, afternoon, and night shifts to capture staff working all three shifts. The questionnaire comprised four sections:

Section A captured demographic data,

Sections B, C, and D addressed healthcare provider competence, provider motivation, and resource availability in relation to the delivery of quality healthcare.

The instrument was reviewed by public health and medical experts and revised in consultation with the study supervisor to establish content validity, and its reliability was assessed through a pilot survey that led to the removal of ambiguous items.

### Eligibility Criteria

All clinical and administrative staff actively working at the hospital – who gave clear informed consent – were eligible for inclusion. Non-clinical support staff (porters, cleaners, mortuary attendants, cooks), students on internship or rotation, visiting healthcare workers, and individuals who declined or withdrew consent were excluded.

### Ethical Considerations

Ethical approval to conduct the study was obtained through the Department of Public Health, University of Makeni, and the Medical Superintendent of Makeni Regional Hospital granted permission. The purpose of the study was explained to all prospective respondents, and informed consent was obtained after assuring participants that their responses would be used solely for academic purposes and treated as strictly confidential. Participation was voluntary throughout.

### Data Analysis

Quantitative data were analyzed using STATA. Descriptive statistics, including frequencies, percentages, means, and standard deviations, were generated to characterize the sample and to summarize responses on competence, motivation, resource availability, and their perceived positive and negative impacts on quality healthcare delivery.

## RESULTS

### Demographic Characteristics

Of the 155 respondents, 61.3% were female and 38.7% were male. More than half (52.3%) were aged 30-39 years, 26.5% were 20-29 years, 14.2% were 40-49 years, and 7.1% were 50 years and above. Slightly more respondents identified as Muslim (53.0%) than Christian (47.1%).

Most respondents (55.5%) had 1-5 years of service, 31.0% had 6-10 years, 7.7% had 11-15 years, and 5.8% had 16 years or more. The majority were married (55.5%), 44.0% were single, and 0.65% were widowed.

Nurses formed the largest professional cadre (66.5%), followed by laboratory scientists/technicians (13.6%); the remaining cadres (CHOs, pharmacists, physician assistants, medical doctors, administrative staff, radiographers, and others) each accounted for less than 5% of respondents. Most respondents (67.7%) held pin-coded (permanent) positions, while 32.3% were volunteers.

**Table 1:**
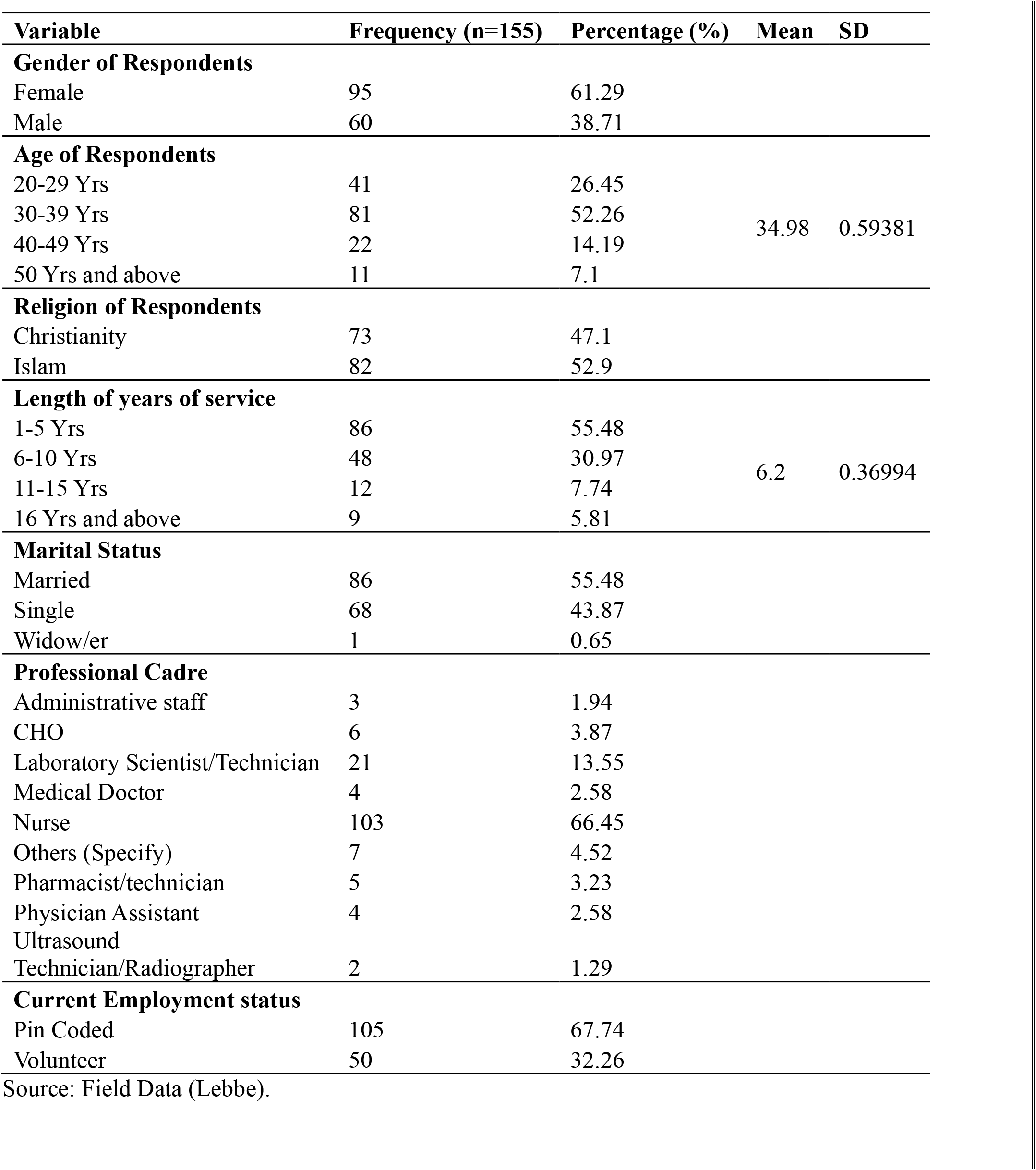
Demographic Characteristics of respondents.

### Service Provider Competence and Quality Healthcare Delivery

Most respondents (74.8%) reported possessing the required professional skills for their role, and 78.1% perceived that their facility was providing quality healthcare.

About two-thirds (67.1%) felt that patients accessed quality health services easily.

Regarding perceived staff competence, 73.6% described their departmental staff as competent, 18.1% as very competent, 1.9% as not competent, and 6.5% did not know.

On the perceived positive impacts of competent personnel, 94.8% associated competence with improved patient outcomes, 91.2% with improved delivery of quality healthcare, 77.42 with effective use of resources, 56.8% with increased service utilization, and 39.4% with self-fulfillment as a provider.

Conversely, on the perceived negative impacts of incompetence, 90.3% associated incompetent staff with poor quality healthcare delivery, 89.7% with poor patient outcomes, 71.7% with inefficient use of resources, and 57.4% with reduced service utilization.

**Table 2.** Service provider competence and quality healthcare delivery (n = 155)

| Variable | Frequency (n=155) | Percentage (%) |
| --- | --- | --- |
| **Personnel have the required professional skills** |  |  |
| No | 39 | 25.16 |
| Yes | 116 | 74.84 |
| **Perceived that the facility is providing quality healthcare** |  |  |
| No | 34 | 21.94 |
| Yes | 121 | 78.06 |
| **Patients access quality health service easily** |  |  |
| No | 51 | 32.9 |
| Yes | 104 | 67.1 |
| **Competency of staff in the facility** |  |  |
| Competent | 114 | 73.55 |
| I don't know | 10 | 6.45 |
| Not competent | 3 | 1.94 |
| Very competent | 28 | 18.06 |
| **Perceived Positive Impacts of Staff Incompetence on Delivery of Quality Healthcare Delivery at the Makeni Regional Hospital** |  |  |
| **Improve patient outcome** |  |  |
| No | 8 | 5.16 |
| Yes | 147 | 94.84 |
| **Effective use of Resources** |  |  |
| No | 35 | 22.58 |
| Yes | 120 | 77.42 |
| **Quality healthcare delivery** |  |  |
| No | 13 | 8.39 |
| Yes | 142 | 91.61 |
| **Increase service utilization** |  |  |
| No | 67 | 43.23 |
| Yes | 88 | 56.77 |
| **Self-fulfilment as a provider** |  |  |
| No | 94 | 60.65 |
| Yes | 61 | 39.35 |
| **Perceived Negative Impacts of Staff Incompetence on Delivery of Quality Healthcare Delivery at the Makeni Regional Hospital** |  |  |
| **Poor patient outcome** |  |  |
| No | 16 | 10.32 |
| Yes | 139 | 89.68 |
| **Inefficient use of resources** |  |  |
| No | 44 | 28.39 |
| Yes | 111 | 71.61 |
| **Poor quality healthcare delivery** |  |  |
| No | 15 | 9.68 |
| Yes | 140 | 90.32 |
| **Reduce service utilization** |  |  |
| No | 66 | 42.58 |
| Yes | 89 | 57.42 |

**Figure 1:**
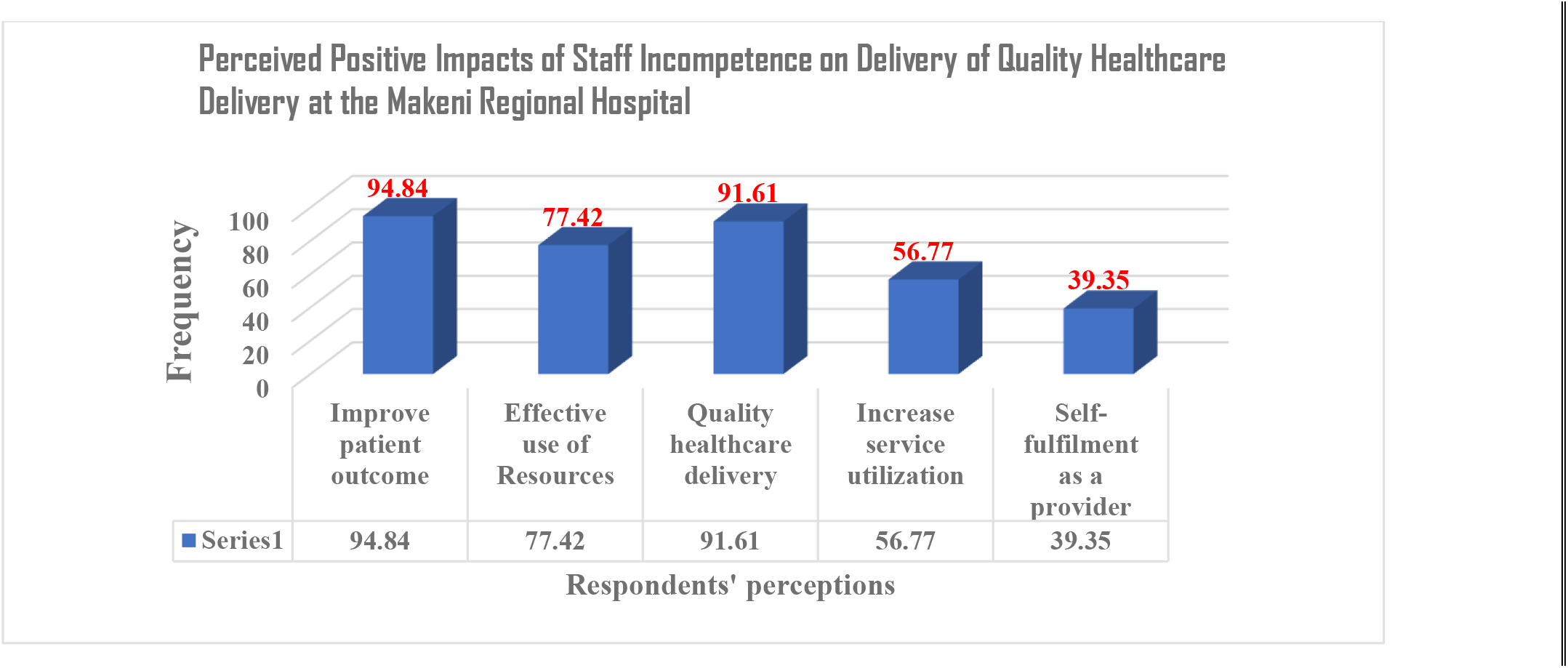
Perceived Positive Impacts of Staff Incompetence on Delivery of Quality Healthcare Delivery at the Makeni Regional Hospital.

**Figure 2:**
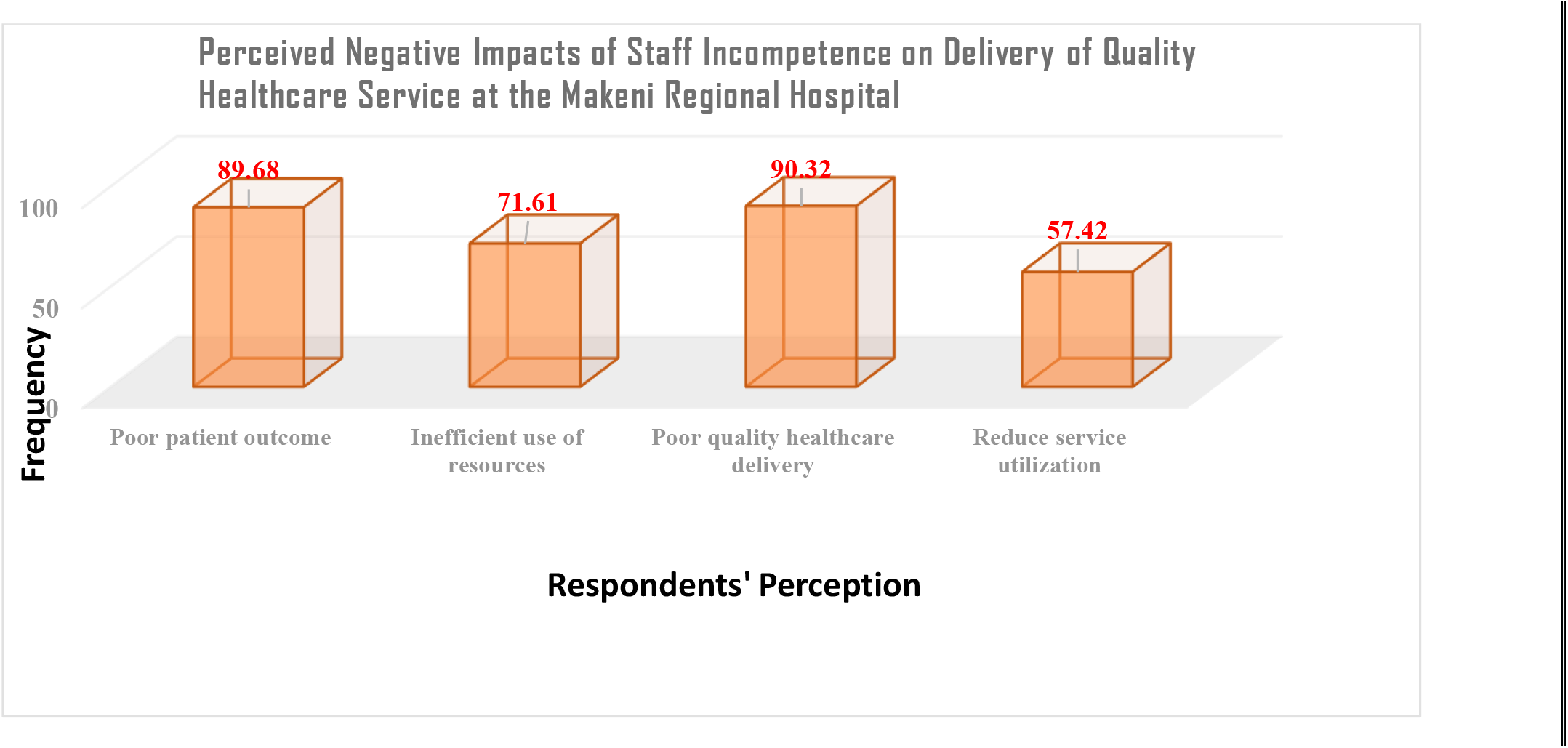
Perceived Negative Impacts of Staff Incompetence on Delivery of Quality Healthcare Service at the Makeni Regional Hospital.

## DISCUSSION -- RESULTS

This study assessed the influence of service provider competence, motivation, and resource availability on quality healthcare delivery at Makeni Regional Hospital.

The high proportion of respondents reporting the required professional skills (74.9%) and perceiving their colleagues as competent (91.7%) combined competent/very competent) aligns with the view that healthcare professionals possess elevated competence given the nature of the services they provide and their central role in improving patient outcomes (Shahrbabaki et al., 2020). This is consistent with the broader literature framing competence as a set of observable, measurable knowledge, skills, and attributes that shape job performance (Kim & Jung, 2022a), and with evidence that abilities, attitudes, and dedication of health professionals are central to hospital performance (Laititi, 2022).

Provider motivation told a markedly different story: only 7.1% of respondents were highly motivated, despite unanimous agreement (100%) that motivation affects quality of care.

The gap between recognized importance and actual motivation is consistent with prior findings that low employee motivation is a significant contributor to poor service quality, linked to absenteeism, irritability with patients, long wait times, informal fee charging, and labor unrest (Alhassan et al., 2013).

It also echoes evidence from rural Sierra Leone that unfavorable working conditions, limited training access, and long hours driven by staffing shortages demotivate health workers (Wurie et al., 2016)

The prominence of finance, accommodation, job satisfaction, and in-service training as cited forms of motivation supports findings that reimbursement and benefit-administration practices are meaningfully correlated with the standard of healthcare delivered (Bula et al., 2018), and that both monetary and non-monetary incentives increase employee motivation and organizational loyalty (Bula et al., 2018).

Resource availability emerged as the determinant most strongly and consistently linked to quality outcomes, with 99.4% of respondents affirming its impact.

This finding is consistent with the view that inadequate infrastructure and a lack of competent labor make it difficult for health organizations to meet demand for care (Mosadeghrad, 2013), and with evidence that subpar equipment and materials reduce employee productivity while resource shortages increase workplace stress among providers (Mosadeghrad, 2014b).

The specific deficits identified in this study, inadequate bed capacity (45.81%) and grossly inadequate information technology infrastructure (40%), point to structural gaps that persist even where human resource inputs (competence) appear comparatively strong, underscoring that human resources are only one of several inputs, alongside consumables and physical capital, that health systems require in tandem (Laititi, 2022).

## DISCUSSION -- IMPACT

Taken together, the pattern of high competence, low motivation, and uneven resource adequacy suggests that the weakest link in quality healthcare delivery at Makeni Regional Hospital is not provider skill but the systems that sustain and equip that skill, namely motivation mechanisms and resource provisioning.

This is consistent with broader observations that even well-developed health systems with available resources continue to show significant variation in care standards (Obina, 2019), and that closing this gap requires attention beyond training alone.

## CONCLUSION

Service provider competence, motivation, and resource availability were each found to significantly influence the delivery of quality healthcare at Makeni Regional Hospital.

Competence and provision of quality health service were closely related, with the hospital’s workforce demonstrating a generally high level of professional skill. However, motivation was low, with only 7.1% of respondents highly motivated, despite universal agreement that motivation shapes the quality of care delivered. This may be paradoxical.

Resource availability was affirmed by 95.5% of respondents as enhancing quality healthcare delivery and was the factor most consistently linked to improved patient outcomes, though notable gaps remain in bed capacity and information technology infrastructure.

## Data Availability

All data produced in the present study are available upon reasonable request to the Corresponding or Lead Authors.

## ABBREVIATIONS

DHS: Demographic Health Survey
FHCI: Free Health Care Initiative
HRIS: Human Resource Information System
LMCs: Low-Middle-Income Countries
MDG: Millennium Development Goal
MoHS: Ministry of Health and Sanitation
MoHSS: Ministry of Health and Social Security

## ACKNOWLEDGEMENTS

All authors thank the data collection team for their well-done job, the Sierra Leone Ministry of Health and Sanitation for giving access to their facility and data for this research study, and the management and administration of the University of Makeni (UNIMAK) for the provision of students who helped with the data collection process.

Also, Lee thanks the U. S. Fulbright Foreign Scholarship Board for a Visiting Fulbright Teaching and Research Scholar Award at the University of Malta for 2025-2026.

## DATA AVAILABILITY

All data for our research study are presented within this article and are available separately from the Lead Author or Corresponding Author with a request from any interested individual.

## ETHICS APPROVAL and CONSENT to PARTICIPATE

Ethical approval was sought and received from the Ethical Review Board of the Sierra Leone Ethics and Scientific Review Committee. An informed consent form was issued to each respondent to seek their consent, assure them that their contribution to the study will be confidential, and that the data provided can only be used for the purpose of the study. The respondents were also informed that no form of financial incentive will be given to them for being part of the study – and that their choice of not being part of the study cannot stop them from getting any benefit that may come because of the study.

## AUTHORS CONTRIBUTION

**Aiah Lebbie:** Conceptualization, Investigation, Initial Analysis, and Drafting and Editing Final Manuscript.

**Abraham I. Jimmy:** Co-Conceptualization, Data Review, Co-Analysis,plus Reviewing & Proofing Final Draft.

**Sallu N. Kamara:** Supervision, Methodology, and Co-Data Review,plus Reviewing & Proofing Final Draft.

**Lee Presley Gary, Jr**.: Resources, Visualization, Restructuring Outline,plus Editing and Finalizing Manuscript.

## AUTHOR BIOGRAPHIES

**Aiah Lebbie** is currently serving as a Pediatric Surgeon at the University of Sierra Leone / Teaching Hospital Complex-Connaught and is the only Pediatric Surgeon in the country. He is a Lecturer with the College of Medicine and Allied Health Sciences at the University of Sierra Leone. Dr. Lebbie travels to the provinces to perform surgeries on children of all ages at no cost to the patients. He was awarded the Medical Practitioner of the Year 2019 by All Walks of Life..

- Sallu Nfagie Kamara is a Lecturer on the Faculty of Pharmaceutical Sciences and Department of Pharmacology at the College of Medicine and Allied Health Sciences with the University of Sierra Leone, He is a graduate of the University of Sierra Leone and earned a Master’s Degree from the University of Lagos (Nigeria), plus a Master of Public Health from the University of Maken (Sierra Leone).
- Abraham Isiaka Jimmy is a Public Health Specialist, Researcher and an University Instructor. He has over seven years of experience working in emergency management and community development. He teaches graduate public health and mental health courses at the University of Makeni, located in Sierra Leone. A l s o, h e s e r v e s a s a n a d v i s e r f o r One Health Program.
- Lee P. Gary, Jr. is as Visiting Fulbright Research Scholar at the University of Malta, and he is the Director of the Maltese Legionella Project. Previously, he was a Visiting Fulbright Fellow with the new School of Public Health at the University of Makeni in Sierra Leone for 2025.

